# Altered brain reward response to monetary incentives in fibromyalgia: A replication study

**DOI:** 10.1101/2022.03.02.22271367

**Authors:** Su Hyoun Park, Eden Z. Deng, Anne K. Baker, Kelly H. MacNiven, Brian Knutson, Katherine T. Martucci

## Abstract

Dysregulated brain reward systems have been observed in chronic pain. Using functional magnetic resonance imaging (fMRI) and a monetary incentive delay (MID) task, Martucci et al. (2018) showed that neural responses to reward anticipation and outcome are altered in patients with fibromyalgia. The current study aimed to replicate these results in a separate cohort of patients with fibromyalgia recruited at a new location using a similar study design. Twenty patients with fibromyalgia and 20 healthy controls were included in the replication study. Group fMRI analyses revealed a solid and consistent trend of main findings similar to the previous results. Specifically, in the replication cohort of patients with fibromyalgia, medial prefrontal cortex (MPFC) activity was reduced during gain anticipation and increased during no-loss (non-punishment) outcomes, as compared to controls. Similar to the Martucci et al. results, again in the replication cohort, nucleus accumbens activity during gain anticipation did not differ in patients compared to controls. The same behavioral, correlational, and exploratory analyses that were conducted in the Martucci et al. study were conducted in the present replication study, with prior results largely replicated here. Thus, the present replication study results solidify observations of altered cortico-striatal processing to monetary rewards in chronic pain, which underscore relevance of altered brain reward circuits, particularly as related to the MPFC in patients with fibromyalgia.

## 1. INTRODUCTION

Brain reward systems play an essential role in modulating pain experiences (Becerra et al., 2001; Becerra & Borsook, 2008; Borsook et al., 2016; DosSantos et al., 2017; Fields, 2006; Leknes & Tracey, 2008). Prior neuroimaging studies provide evidence of brain reward/valuation system alterations in patients with chronic pain (Baliki et al., 2010, 2012; Berger et al., 2014; DosSantos et al., 2017; Martucci et al., 2018, 2019; Park et al., *preprint*). As these alterations may reflect dysregulated reward behavior and possibly be used as a biomarker of pain chronification (Baliki et al., 2012), it is crucial to understand and verify altered brain rewards systems in chronic pain and their relation to subjective pain experiences.

Reward processing in the human brain involves the cortico-subcortical brain networks, including the nucleus accumbens (NAcc), medial prefrontal cortex (MPFC), ventral tegmental area (VTA), anterior insula (aINS), and anterior cingulate cortex (ACC) (Haber & Calzavara, 2009; Haber & Knutson, 2010; Sescousse et al., 2013). These areas are not only strongly implicated in reward processing (Bush et al., 2002; Haber & Calzavara, 2009; Knutson et al., 2001; Preuschoff et al., 2008) but also known to send direct projections to each other (Chikama et al., 1997; Fallon, 1981; Fiorillo et al., 2003; Kunishio & Haber, 1994).

Alterations of brain reward systems in chronic pain are shown in many pain conditions, including patients with chronic back pain and fibromyalgia. Fibromyalgia is a condition of widespread chronic pain typically accompanied by symptoms of cognitive deficits, fatigue, and mood disorders such as anxiety and depression (Sluka & Clauw, 2016). Compared to healthy controls, reduced responses in the right striatum to reward stimuli were observed in patients with chronic low back pain and fibromyalgia (Kim et al., 2020). Also, greater NAcc-MPFC functional connectivity was shown in chronic back pain patients than healthy controls when receiving heat stimulation and performing a self-report pain-monitoring task (Baliki et al., 2010). Several neuroimaging studies revealed dysregulated brain reward systems in fibromyalgia. For example, reduced midbrain activity was observed in patients with fibromyalgia during rewarding event/punishment anticipation (i.e., pain relief/anticipation) (Loggia et al., 2014).

Using functional magnetic resonance imaging (fMRI), Martucci et al. (2018) found that neural responses to monetary rewards are altered in patients with fibromyalgia, such that patients showed reduced MPFC activity during anticipation of gains, but greater activation in the MPFC after avoiding losses compared to a healthy population. Moreover, NAcc activity during reward anticipation showed no significant group differences. Patients with fibromyalgia also showed slightly reduced activity in the VTA, aINS, and ACC during reward anticipation compared to healthy controls.

The aim of this study is to closely examine the replicability of the results of Martucci et al. (2018) in a separate cohort of patients with fibromyalgia. As outlined in our pre-registered plan on the Open Science Framework (OSF, https://osf.io/4yctn), we hypothesized that similar to results from Martucci et al. (2018), altered brain reward activity in patients with fibromyalgia would be observed compared to healthy controls. Specifically, patients would show reduced MPFC activity during gain anticipation but greater MPFC activity after avoidance of loss outcomes compared to healthy controls. NAcc activity during reward anticipation, in contrast, should show similar responses in both groups. We also conducted correlation and exploratory analyses previously performed by Martucci et al. (2018). We specifically examined whether the findings of correlations between several clinical measures and brain regions would reveal similar results with Martucci et al. (2018) and compared extracted fits of VTA, aINS, and ACC activity between patient and control groups during reward anticipation.

## 2. METHODS

### 2.1. Participants

Twenty individuals diagnosed with fibromyalgia and 20 healthy individuals participated in the study. Patients with fibromyalgia were required to 1) meet modified American College of Rheumatology 2016 criteria for fibromyalgia (Wolfe et al., 2016), 2) have pain in all four quadrants of the body, and 3) have an average pain score of at least 2 (0–10 verbal scale) over the previous month. All patients had no uncontrolled depression or anxiety and did not take any opioid medications. All study procedures were approved by the Duke University Institutional Review Board.

### 2.2. Medication usage

All controls reported taking no pain or mood-altering medications. All patients were opioid naïve (i.e., less than one month of opioid use within their lifetime and no opioid use before the study for > 90 days). Sixteen of the patients were taking nonsteroidal anti-inflammatory drugs (NSAIDs, N = 3), acetaminophen (N = 2), serotonin-norepinephrine reuptake inhibitors (SNRIs, N = 7), selective serotonin reuptake inhibitors (SSRIs, N = 5), tricyclic antidepressants (TCAs, N = 3), other anxiolytics (e.g., buspirone hydrochloride, N=1), triptans (N = 1), trazodone (N = 1), antiepileptic (N = 3), muscle relaxants (N = 5), gamma-aminobutyric acid (GABA) analogs (e.g., pregabalin and gabapentin, N = 4), benzodiazepine (N = 3), norepinephrine–dopamine reuptake inhibitor (NDRI, N = 3). The four remaining patients reported not taking any pain or mood-altering medications.

### 2.3. Study Procedures

All study procedures were conducted at the Duke-UNC Brain Imaging and Analysis Center (BIAC). All participants received the instructions of the monetary incentive delay (MID) and the arousal and valence rating tasks and practiced them before scanning. In addition, patients with fibromyalgia and healthy controls completed questionnaires including Beck Depression Inventory (BDI; Beck et al., 1988), State-Trait Anxiety Inventory (STAI-State, STAI-Trait; Spielberger et al., 1970), Behavioral Inhibition System/Behavioral Approach System (BIS/BAS Scales; Carver & White, 1994), Profile of Mood States (POMS; McNair et al., 1971), Positive and Negative Affect Schedule (PANAS; Watson et al., 1988), Brief Pain Inventory (BPI; Keller et al., 2004), PROMIS Fatigue (Cella et al., 2010), and Brief Symptom Inventory (BSI; Derogatis & Melisaratos, 1983).

### 2.4. Monetary Incentive Delay (MID) Task

The task was programmed in MATLAB with Psychophysics Toolbox v.3 (Brainard, 1997; Kleiner et al., 2007), and the procedures were the same as the procedures used in Martucci et al. (2018). All participants performed two runs of the MID task. Each trial consisted of 1) a cue, 2) a fixation cross, 3) a triangle target, and 4) feedback. A cue denoted the amount of money that participants can either gain (+$5, +$1, +$0, accompanied with a “circle” symbol) or lose (-$5, - $1, -$0, accompanied with a “square” symbol). After each cue, a fixation mark was presented for two seconds, and then a triangle target was presented. An initial target duration was set between 260 ms∼280 ms, and the target was presented for variable durations. Based on the participant’s performance, the target duration was automatically adjusted to achieve a hit rate of approximately 66% across trials. Participants were instructed to press the button as quickly as possible. Then, feedback was given. If participants’ response resulted in a “hit” (i.e., press the button before target offset) on gain trials, they won that amount of money, but if their response resulted in a “miss” (i.e., no button press or pressing the button after target offset), they did not win any money. If they “hit” on loss trials, they avoided losing that amount of money, but in the case of a “miss,” they lost that amount of money (Fig. 1). After the MID task functional scans, all participants completed the rating (i.e., arousal, valence) task. Participants were instructed to rate levels of valence and arousal for each cue that they saw during the MID task (e.g., +5 accompanied with a circle cue). For this rating, 7-point Likert scales were presented on a screen, and participants rated via button press the valence (negative – neutral – positive) and arousal (low – medium – high) scales.

**Figure 1.**
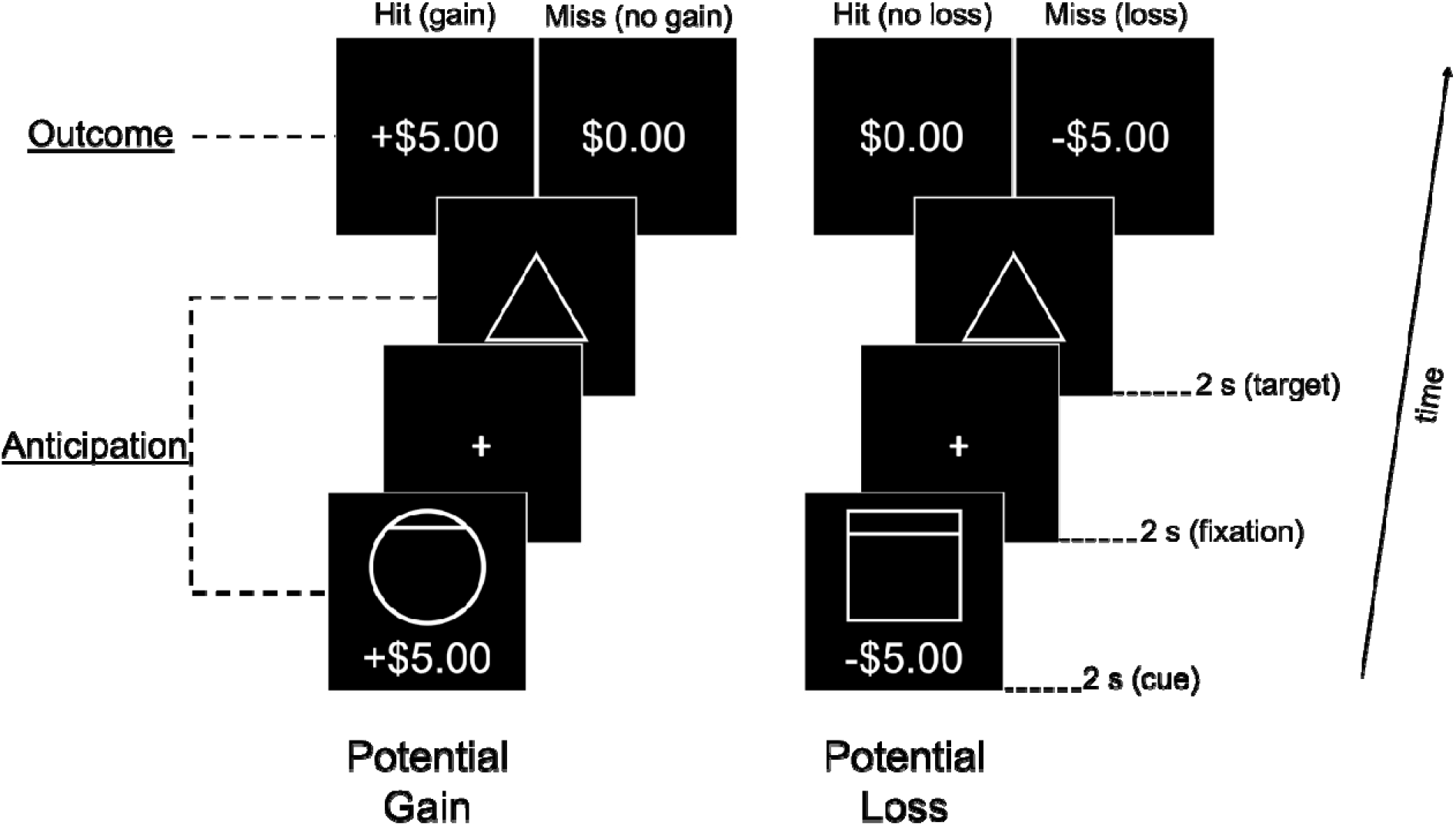
The general procedure of monetary incentive delay (MID) task. Each trial consisted of anticipation and outcome phases, and this example depicts both gain and loss trials. Consistent with Martucci et al. (2018), each trial (TR-locked; TR=2 seconds) consisted of TR 1 = cue, TR 2 = fixation, TR 3 = target, and TR 4 = feedback, TR(s) 5∼7 = variable duration inter-trial interval. Cues were either circles (potential gains) or squares (potential losses), and they were presented with monetary values. After a fixation period, a target period began. A triangle was presented for a variable duration (∼250 ms) during the target period, which was determined on prior response accuracy to obtain an average 66% hit rate. During the outcome phase, win or loss (i.e., hit or miss) feedback was given. After the feedback, a black screen was presented as a pseudo-randomized inter-trial interval period (2, 4, or 6 s duration).

**Figure 1.**
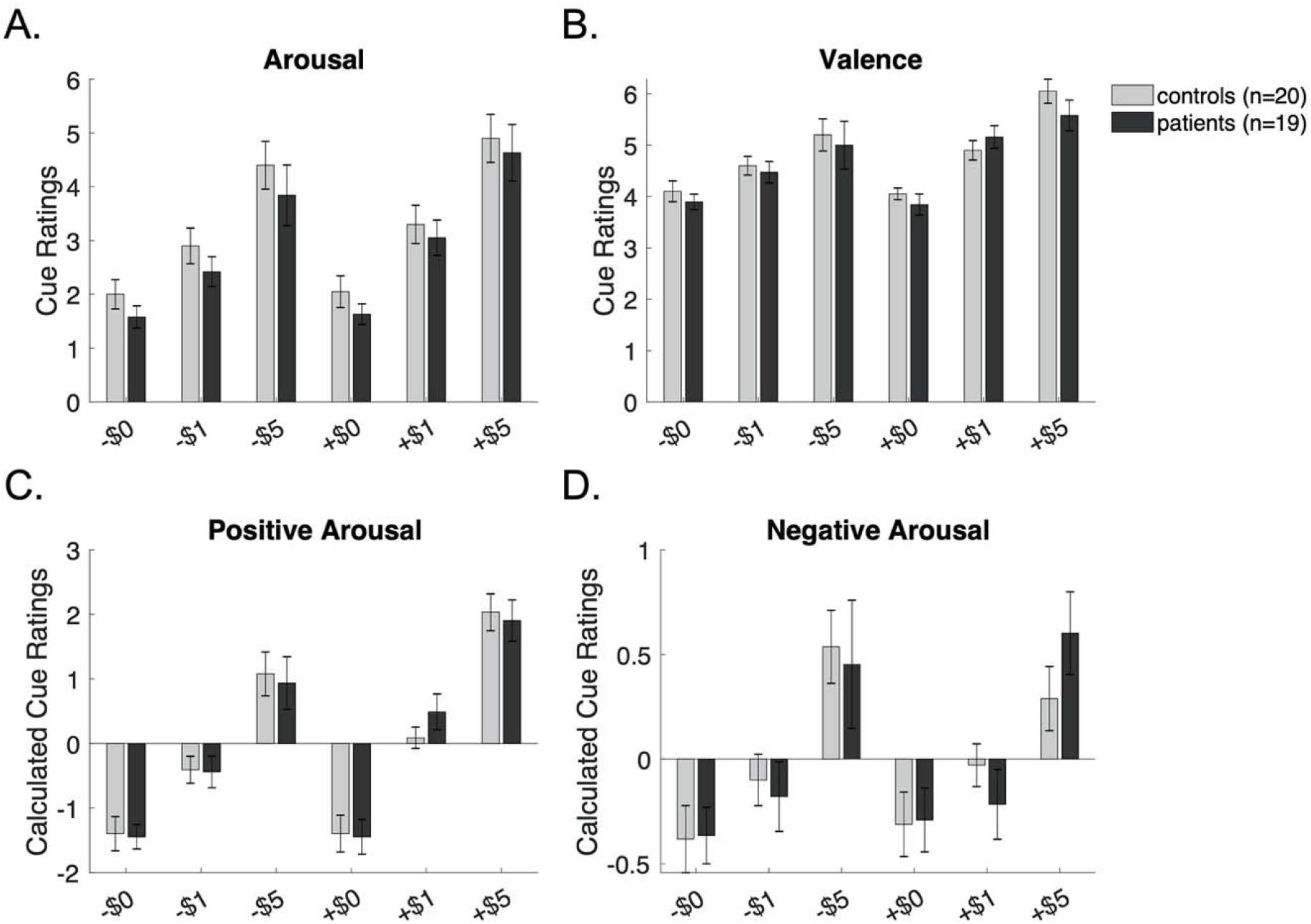
Cue Ratings of Arousal, Valence, Positive Arousal, Negative Arousal. In Martucci et al. (2018), arousal ratings were lower in the patient group compared to the control group across all cues (but other ratings were similar in both groups). In our study, (A) arousal, (B) valence, (C) positive arousal, and (D) negative arousal ratings showed no differences in patients and controls. One patient did not provide any ratings so was not included in the analyses.

### 2.5. MRI Scans

Scans were conducted on a 3T GE Premier UHP system with a 48-channel coil at the Duke-UNC Brain Imaging and Analysis Center. Consistent with Martucci et al. (2018), the scan sessions consisted of the initial preparatory localizer, asset calibration scans, two MID task fMRI scans, and T1 anatomical scan. The fMRI scan parameters were as follows: Gradient Echo Pulse Sequence, flip angle 77°, echo time (TE) 25 ms, repetition time (TR) 2 seconds, sequential descending slice order, 46 slices, 2.9 mm slice thickness, pixel size 2.9 mm. Unlike Martucci et al. (2018), we did not use a Spiral In-Out scan sequence. The 2 MID task scans consisted of 266 and 302 volumes, respectively, and we excluded the first 6 lead-in volumes (12 s) and 4 lead-out volumes (8 s). A T1 anatomical scan (MPRAGE sequence) was acquired for registration of functional images with parameters as follows: whole-brain coverage including the brainstem and cerebellum, 1 mm slice thickness, 256 mm frequency field of view (FOV), frequency direction anterior/posterior, flip angle 8°, TR 2.2 seconds, TE 3.2 ms.

### 2.6. MID fMRI Scan: Preprocessing

All neuroimaging data were preprocessed using Python and AFNI software (Analysis of Functional Neuroimages). For preprocessing, the following GitHub repository was used: https://github.com/kellyhennigan/MID_processing_example. Specifically, custom scripts were used for functional images to remove the first 6 lead-in volumes and perform slice time correction and motion correction. Next, images were spatially smoothed with a 4 mm (full-width half-maximum, FWHM) Gaussian kernel and converted to be in units of percent change values (i.e., normalization). High-pass filters (a threshold of 0.011 Hz) were applied to remove low-frequency fluctuations. Then, the concatenated preprocessed functional images were transformed to standard group space. Anatomical images were skull stripped (3dSkullStrip) and aligned to Talairach space (AFNI’s @auto_tlrc). Next, the first volume of the functional image was co-registered to the anatomical image, and then this volume was skull stripped (3dSkullStrip). Lastly, the anatomical image was co-registered to functional data and transformed from native to standard space.

### 2.7. fMRI Task Contrasts

Four orthogonal regressors were modeled to contrast participants’ responses to gain and loss vs. neutral trials (i.e., no-gain or no-loss) during anticipation and outcome phases. The use of these regressors resulted in four contrasts: gain vs. no-gain anticipation (GVNant), loss vs. no-loss anticipation (LVNant), gain vs. no-gain outcome (GVNout), and no-loss vs. loss outcome (NVLout). However, based on findings in Martucci et al. (2018), our main analysis in the present study focused on the following two contrasts (planned in our pre-registered analysis in OSF, https://osf.io/4yctn):

1. gain versus no-gain anticipation (GVNant): gain (+$5) trials versus no-gain (+/−$0) trials during the anticipation period
2. no-loss versus loss outcome (NVLout): hits (−$0 outcome) versus misses (−$5 outcome) during the outcome period

The regressors were convolved with a single-gamma hemodynamic response function, with approximately 6-second delay. Motion censoring was applied to censor volumes showing motion greater than a euclidean norm value of 0.5.

### 2.8. Region of Interest Analyses

Consistent with Martucci et al. (2018), primary region of interest (ROI) analyses were focused on the NAcc and MPFC areas. We used the same ROI masks that were used in Martucci et al. (2018). Specifically, the bilateral NAcc ROI was created from the DKD_Desai_MPM atlas included in AFNI. The MPFC ROI was fixed 8-mm diameter spheres centered at +/−4, 50, −3 (see Martucci et al., 2018 for detailed information for creating these ROIs).

### 2.9. Statistical Analysis

Statistical analyses strictly followed those reported in Martucci et al. (2018). For behavior data, group and between-group level statistics for participants’ reaction time and accuracy rates during the MID task were measured. After performing the MID task, participants were asked to rate their arousal and valence responses to each cue that appeared during the MID task (e.g., a rectangle cue accompanied with -$5). Participants’ arousal and valence responses to each cue were measured. Based on these ratings, participants’ positive and negative arousal were also calculated (i.e., mean-deviated and rotated) as performed in Martucci et al. (2018). Two-way ANOVA (Group: patient vs. controls, and cue: +/− $0, $1, $5) was conducted for arousal, valence, positive arousal, and negative arousal. Due to missing data, one patient was excluded from analyses of cue ratings.

For fMRI data, group and between-group level statistics for averaged beta values for each ROI and contrast combination were conducted. The same custom MATLAB scripts used in Martucci et al. (2018) were used to extract these values, and as used in Martucci et al. (2018) and planned on the pre-registration, the significance threshold was set at p < 0.0125. For visualization purposes, AFNI’s 3dttest++ was used to create activation maps, and ROI group results were thresholded at a voxelwise threshold of p < 0.05.

Correlational analyses were conducted between fMRI beta values and questionnaire measures. Consistent with Martucci et al. (2018), correlations were tested for combined patient and control groups using JASP (JASP Team, 2020). ROI fMRI beta values were also restricted to NAcc GVNant, MPFC GVNant, and MPFC NVLout (i.e., three independent measures for correlations between ROI fMRI beta measures). Clinical measures included three BAS subscales (behavioral drive, behavioral reward responsiveness, and behavioral fun seeking), behavioral inhibition (BIS subscale), positive affect (PANAS, PAS subscale), negative affect (PANAS, NAS subscale), total mood disturbance (POMS), depression (BDI), trait anxiety (STAI Trait), state anxiety (STAI State), pain severity (BPI), pain interference (BPI), fatigue (PROMIS Fatigue), and the depression and anxiety subscales of the Brief Symptom Inventory (BSI). Correlations identified among clinical measures revealed 3 independent clusters of measures: 1) BAS fun and BAS reward (p < 0.001), 2) BAS drive and BIS (p < 0.001), and 3) all others (p < 0.025). Thus, a total of 6 independent clusters of measures were identified, and accordingly, the ROI fMRI beta vs. clinical measures correlational analyses were Bonferroni corrected for a total of 6 independent comparisons and determined to be significant at the level of p < 0.008 (corrected threshold). As set up in Martucci et al. (2018), post-hoc analysis for within-group (i.e., separately for patients and controls) was conducted if the initial across group analyses showed significant correlations (p < 0.008). All results are reported in Supplementary Table 1.

### 2.10. Additional ROIs and Contrasts for Post-hoc Analysis

As conducted in Martucci et al. (2018), we implemented exploratory analyses and ran additional analyses using the VTA, aINS, and ACC ROIs. For these exploratory analyses, we used the same ROI masks that were used in Martucci et al. (2018). All ROIs were resampled to functional image dimensions (using AFNI’s 3dfractionize, clip 0.1) and transformed back to Talairach space (using AFNI’s adwarp; see Martucci et al., 2018 for detailed information on creating these ROIs).

Additionally, as exploratory analyses, we added two task contrasts, 1) loss versus no-loss anticipation (LVNant): loss (−$5) trials versus no-loss (+/−$0) trials during the anticipation period, and 2) gain versus no-gain outcome (GVNout): hits (+$5 outcome) versus misses (+$0 outcome) during the outcome period, and ran the NAcc and MPFC ROI analyses. Martucci et al. (2018) did not include the +$1 and −$1 trials in any analyses based on previous research’s findings of less pronounced brain responses in smaller incentives (Knutson et al., 2003, 2005). For exploratory analysis, we added -$1 trials to our primary analysis of an MPFC NVLout condition, which only consisted of -$5 trials (i.e., hits [−$0 outcome] versus misses [−$5 outcome] during the outcome period). We found that, as compared to anticipation trials, the number of outcome trials in each condition (i.e., -$5 and -$1/hit and miss) varied, which might yield less power; thus, as an exploratory analysis, we included -$1 trials for the NVLout condition to increase the power.

### 2.11. Whole Brain and Expanded Anterior Medial Prefrontal Cortex Mask Post-hoc Analysis

Consistent with Martucci et al. (2018), post-hoc whole brain and large anterior MPFC region analyses were conducted in the present study to confirm the ROI findings and compare them with results from parallel analyses in Martucci et al. (2018). Again, we used the same anterior medial prefrontal cortex mask used in Martucci et al. (2018). For visualization purposes, results were thresholded at p < 0.05 (z = 1.975). AFNI’s Clustsim algorithm was used, and cluster correction was set at a minimum cluster size of 20 voxels and used first-nearest neighbor clustering, which requires all voxels to touch a face for inclusion in a cluster (NN set to = 1).

## 3. RESULTS

### 3.1. Clinical, Behavioral, and Psychological Measures

Twenty patients and 20 healthy controls were included in the final analysis (Table 1). The results of clinical, behavioral, and psychological measures were similar to Martucci et al. (2018), except for BIS score (Table 2). Significant differences in measures including mood, fatigue, anxiety, depression, number of pain areas, pain severity, and pain interference were shown between patients and controls, but BIS scores did not differ between groups. As found in Martucci et al. (2018), patients did not show severe levels of depression (BDI score) or anxiety (STAI score).

**Table 1.** Participant Demographics. A total of 40 participants (patients N=20, controls N=20) completed the study. No participants were of race categories for Asian, Pacific Islander, Alaskan, or Native American. “Other” refers to self-identified race other than all of these categories.

|  | **Patients** | **Controls** |
| --- | --- | --- |
| **Total Participants**<br>**(all female)** | **20** | **20** |
| **Righthanded** | **19** | **19** |
| **Self-Identified Race** |  |  |
| **American Indian** | **0** | **1** |
| **Caucasian** | **15** | **17** |
| **African American** | **4** | **2** |
| **Other** | **1** | **0** |
| **Hispanic or Latina**<br>**Ethnicity** | **1** | **1** |
| **Employment Status** |  |  |
| **Part-time employed** | **1** | **1** |
| **Full-time employed** | **14** | **12** |
| **Unemployed** | **5** | **7** |
| **Income Level** |  |  |
| **\$0–\$34,999** | **7** | **3** |
| **\$35,000–\$59,999** | **5** | **4** |
| **\$60,000 or more** | **8** | **11** |

| Education Level |  |  |
| --- | --- | --- |
| High School | 1 | 0 |
| College/University | 13 | 13 |
| Advanced Degree | 6 | 7 |

**Table 2.** Clinical, Behavioral, and Psychological Measures. Not all patients and controls completed all BDI, STAI-State, and STAI-Trait questionnaires. Thus, the total number of participants for each of these measures differs from other measures. Abbreviations: PANAS, Positive and Negative Affect Schedule; BAS, Behavioral Activation System; BIS, Behavioral Inhibition System; POMS, Profile of Mood States; PROMIS, Patient-Reported Outcomes Measurement Information System; BDI, Beck Depression Inventory; FAF, Fibromyalgia Assessment Form; State-Trait Anxiety Inventory (STAI-State, STAI-Trait); BPI, Brief Pain Inventory; BSI, Brief Symptom Inventory; sd, standard deviation. The presented significance values (P-Value) are not corrected for multiple comparisons as results are provided only for descriptive purposes.

|  | Patients |  | Controls |  | P-Value |
| --- | --- | --- | --- | --- | --- |
|  | N | Mean±sd | N | Mean±sd |  |
| Age | 20 | 35.9±12.37 | 20 | 44.25±12.1 | 0.037 |
| Positive Affect (PANAS) | 20 | 24.8±5.7 | 20 | 34.4±7.9 | < 0.001 |
| Negative Affect (PANAS) | 20 | 19.05±5.1 | 20 | 15.15±4.9 | 0.018 |
| Behavioral Drive (BAS) | 20 | 10.8±1.8 | 20 | 11.3±2.3 | 0.458 |
| Behavioral Fun (BAS) | 20 | 10.9±1.9 | 20 | 11.65±2 | 0.246 |
| Behavioral Reward (BAS) | 20 | 17.3±2.4 | 20 | 17.05±2.8 | 0.767 |
| Behavioral Inhibition (BIS) | 20 | 22.3±3.8 | 20 | 20.4±4.6 | 0.17 |
| Total Mood Disturbance (POMS) | 20 | 20±18.4 | 20 | -1.15±8.2 | < 0.001 |
| Fatigue (PROMIS) | 20 | 65.5±6.3 | 20 | 43.6±8.5 | < 0.001 |
| Depression (BDI) | 19 | 16.7±8.7 | 19 | 4.5±6.6 | < 0.001 |
| Number of Pain Areas (FAF) | 20 | 12.05±3.8 | 20 | 0.35±0.58 | < 0.001 |
| **Trait Anxiety (STAI)** | 19 | 48.2±10.4 | 20 | 37.25±9.9 | 0.0018 |
| **State Anxiety (STAI)** | 20 | 38.7±9.3 | 19 | 30.7±7.2 | 0.004 |
| **Pain Severity (BPI)** | 20 | 4.5±1.9 | 20 | 0 | < 0.001 |
| **Pain Interference (BPI)** | 20 | 6.1±2.8 | 20 | 0 | < 0.001 |
| **Brief Symptom Inventory (BSI)**<br>**Depression** | 20 | 5.15±3.6 | 20 | 1.6±2.7 | 0.001 |
| **Brief Symptom Inventory (BSI)**<br>**Anxiety** | 20 | 4.4±3.9 | 20 | 1.7±1.7 | 0.007 |

The number of painful areas per patient and patients’ reported painful regions are shown in Supplementary Fig.1.

Unlike Martucci et al. (2018), in the present study, there was a significant difference in ages between patient and control groups, such that patients were younger than healthy controls. We tried to minimize the age differences during recruitment, but due to challenges with recruiting patients (particularly due to the Covid-19 pandemic), we could not precisely match age between groups. In our primary analyses, we reported results of a one□way ANCOVA, including age as a continuous covariate. The duration of pain related to fibromyalgia symptoms reported by patients ranged from 9 months to 20 years (M = 6.03 years, SD = 5.33 years).

### 3.2. Reaction Times and Accuracy Rates

Similar to Martucci et al. (2018), higher gain (+$5 anticipation) and loss (−$5 anticipation) trials elicited shorter reaction times [F(5,190)=4.2, p = 0.001] for both groups. No group [F(1,190)=0.0, p = 0.842] or group by trial interaction effects [F(5,190)=1.5, p = 0.186] were shown. Consistent with Martucci et al. (2018), we used a version of the MID task that was programmed to track performance and target 66% accuracy rates. Even with this manipulation, again, percent hits were greater for gain and loss trials compared to no gain/no loss trials [F(5,190)=5.3, p < 0.000]. No group [F(1,190)=0.0, p = 0.844] or group by trial interaction effects [F(5,190)=1.7, p = 0.129] were observed. These behavioral results suggest similar engagement and performance across patients (N=20) and controls (N=20).

### 3.3. Cue Ratings: Arousal, Valence, Positive Arousal, Negative Arousal

#### 3.3.1. Arousal

A two-way ANOVA (group x cue) for arousal ratings revealed no significant main effect of group [F(1,185)=1.1, p = 0.301], indicating similar arousal ratings between patients (N=19) and controls (N=20). A significant main effect of cue [F(5,185)=37.3, p = 0.000] was observed for arousal, but no group by cue interaction was observed [F(5,185)=0.1, p = 0.993] (Fig. 2A).

**Figure 2.**
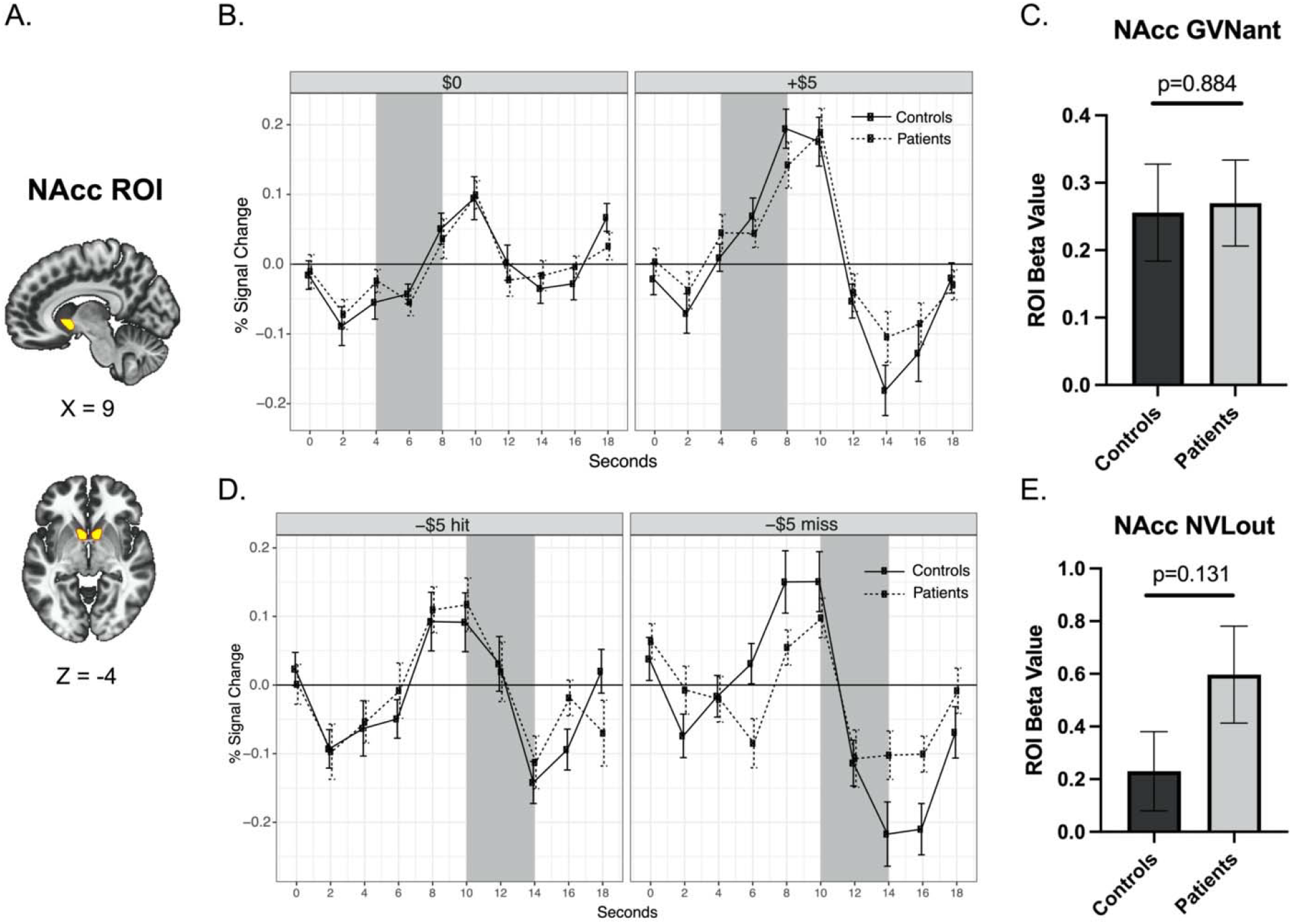
Nucleus Accumbens Activity during Reward Anticipation. (A) Bilateral ROI of the NAcc. The same ROI mask that was used in Martucci et al. (2018) was used in the present analyses. (B) Group means and standard error of raw time course plots of NAcc ROI activity to $0 and +$5 anticipation trials. As plotted in Martucci et al. (2018), the shaded area represents the estimated anticipation period (presentation of cue and fixation [0–4 seconds] plus 4 seconds to account for hemodynamic response function delay). (C) Contrast beta values extracted from the bilateral NACC ROI during reward anticipation (GVNant, +$5 > $0 trials). (D) Group means and standard error of raw time course plots of NAcc ROI activity to -$5 hit (i.e., no loss) and -$5 miss (i.e., loss) trials. (E) Contrast beta values extracted from the bilateral NACC ROI during outcome (NVLout; -$5 hit vs. miss). All beta values are shown as 10^−3^. Abbreviation: NAcc, nucleus accombens; ROI, region of interest.

#### 3.3.2. Valence

A prevalence of positive valence scores for negative cue values indicated potential confusion among participants regarding the instructions for valence ratings. Therefore, valence ratings and negative arousal in this study should be interpreted with caution. A two-way ANOVA (group x cue) for valence ratings revealed no significant main effect of group [F(1,185)=0.7, p = 0.425], indicating similar valence ratings between patients (N=19) and controls (N=20). A cue effect [F(5,185)=20.8, p = 0.000] was observed for valence but no interaction was observed [F(5,185)=0.6, p = 0.732] (Fig. 2B).

#### 3.3.3. Positive Arousal

A two-way ANOVA (group x cue) for positive arousal ratings revealed no effect of group [F(1,185)=0.8, p = 0.367], indicating similar positive arousal ratings in patients (N=19) and controls (N=20). A cue effect [F(5,185)=39.6, p = 0.000] was observed for positive arousal, but no interaction was observed [F(5,185)=0.2, p = 0.954] (Fig. 2C).

#### 3.3.4. Negative Arousal

A two-way ANOVA (group x cue) for negative arousal ratings revealed no effect of group [F(1,185)=1.1, p = 0.306], indicating similar negative arousal ratings in patients (N=19) and controls (N=20). A cue effect [F(5,185)=8.0, p = 0.000] was observed for negative arousal, but no group by cue interaction was observed [F(5,185)=0.4, p = 0.838] (Fig. 2D).

### 3.4. ROI Activation: Nucleus Accumbens

Consistent with Martucci et al. (2018), robust NAcc activity during gain anticipation (GVNant) was observed for both patient and control groups, but no group difference was observed [p=0.884] (Fig. 3). We ran a correlational analysis with NAcc GVNant fMRI beta values and clinical measures (across patient and control groups, N = 40), but unlike Martucci et al. (2018), no significant results were revealed (all p > 0.07). In addition, consistent with Martucci et al. (2018), no significant group differences in NAcc activity during no-loss outcome (NVLout) were observed (Fig. 3). An ANCOVA analysis revealed that the covariate, age, was not significantly related to the NAcc GVNant fMRI beta values, F(1,37)=2.9, p=0.097. There was also no significant group difference in the NAcc GVNant fMRI beta values after controlling for the effect of the age, F(1,37)=0.177, p=0.677. Post-hoc analyses also showed no significant group differences in NAcc activity during loss anticipation (LVNant), and gain outcome (GVNout) (Supplementary Fig. 2).

**Figure 3.**
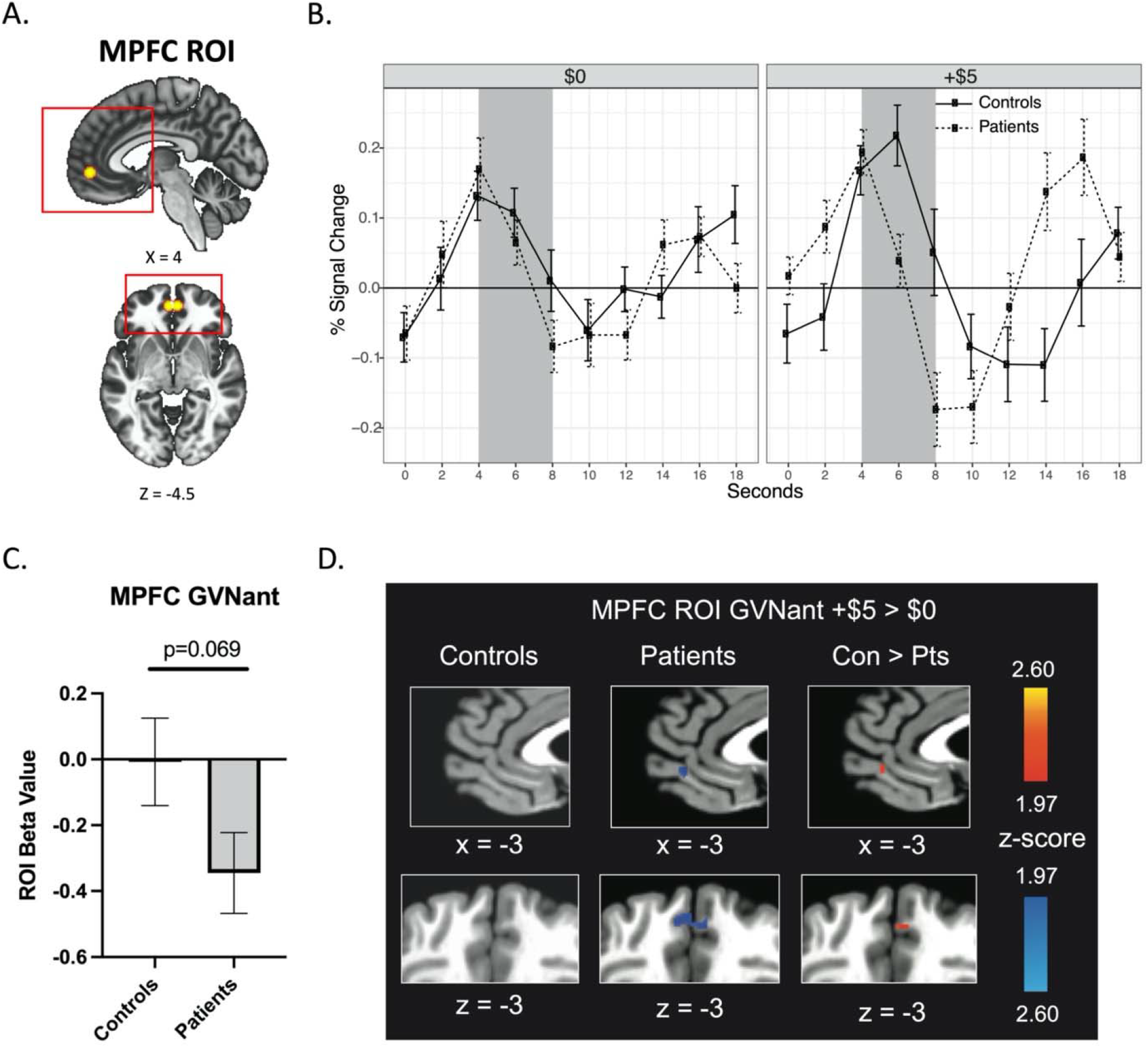
Medial Prefrontal Cortex Activity during Reward Anticipation. (A) Bilateral ROI of MPFC. The same ROI mask from Martucci et al. (2018) was used. Red boxes denote magnification areas of sagittal and axial plane images depicted in Fig 4D. (B) Group means and standard error of raw time course plots of MPFC ROI activity to $0 and +$5 anticipation trials. As plotted in Martucci et al. (2018), the shaded area represents the estimated anticipation period (presentation of cue and fixation [0–4 seconds] plus 4 seconds to account for hemodynamic response function delay). (C) Contrast beta values extracted from the MPFC ROI during reward anticipation (GVNant, +$5 > $0 trials). (D) Contrast activation maps of MPFC ROI activity during reward anticipation (GVNant; p < 0.05, uncorrected). All beta values are shown as 10^−3^. Abbreviation: MPFC, medial prefrontal cortex; ROI, region of interest.

### 3.5. ROI Activation: Medial Prefrontal Cortex

Increased MPFC activity during gain anticipation (GVNant) was observed in the control group relative to the patient group in Martucci et al. (2018). In the current data, no activity was observed in the control group during gain anticipation, but reduced MPFC activity was observed in patients, indicating less MPFC BOLD activity in patients relative to controls (Fig. 4). Even though this result did not reach the corrected statistical threshold of p < 0.0125, the trend was similar to findings of Martucci et al. (2018) (i.e., reduced MPFC activity in patients compared to controls). Possibly due to the reduction in this effect, correlational analyses between the MPFC GVNant fMRI beta values and clinical measures did not reveal significant results. An ANCOVA analysis revealed that the covariate, age, was not significantly related to the MPFC GVNant fMRI beta values, F(1,37) = 0.009, p = 0.923. There was also no significant group difference in the MPFC GVNant fMRI beta values after controlling for the effect of the age, F(1,37) = 3.142, p = 0.085.

**Figure 4.**
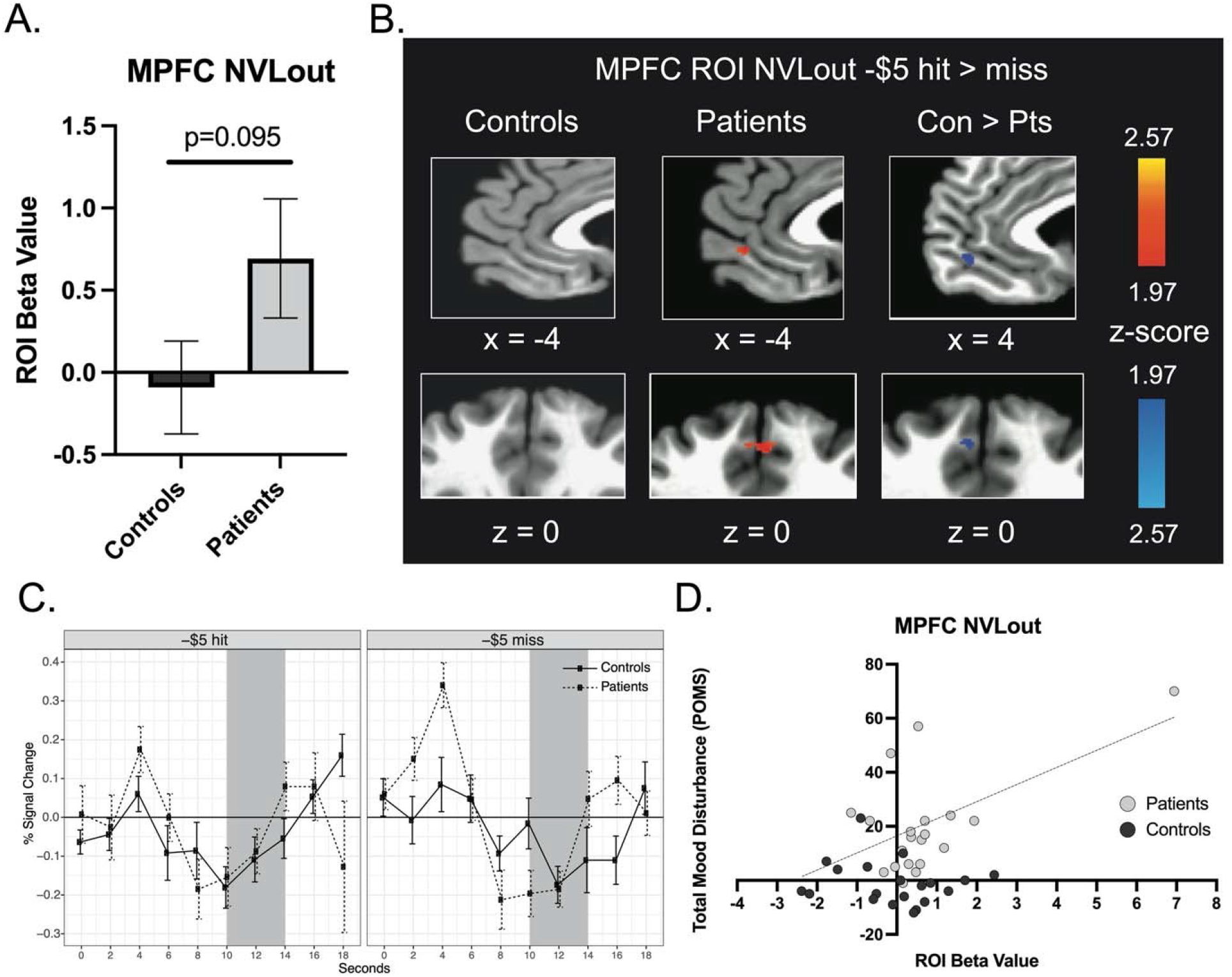
Medial Prefrontal Cortex Activity in Response to Reward No-Loss Outcomes. (A) Contrast beta values extracted from the MPFC ROI during reward outcome (NVLout, -$5 hit > miss). (B) Contrast activation maps of MPFC ROI activity during reward outcome (p < 0.05, uncorrected). (C) Group means and standard error of raw time course plots of MPFC ROI activity to outcomes (hits or misses) for -$5 anticipation trials. As plotted in Martucci et al. (2018), the shaded area represents the estimated outcome period (presentation of outcome and post-outcome [6–10 seconds] plus 4 seconds to account for hemodynamic response function delay). (D) Correlations between MPFC NVLout extracted beta values and total mood disturbance (POMS) for both groups. One patient with a high MPFC NVLout beta value was kept in the analysis because the fMRI scan volumes were censored for bad movements (greater than a euclidean norm value of 0.5) during preprocessing, and only 4 volumes (among 540 volumes) were censored for this patient. Also, this patient did not have as high beta value in the NVLout contrast with both -$5 and -$1 data (see results in Supplementary Figure 3). Therefore, we believe this to be a true representative signal of MPFC activity of -$5 NVLout contrast from this patient. Further, the greater total mood disturbance (worse) in this patient aligns with greater MPFC activation that may be consistent with a greater “relief” experience. Post-hoc analyses excluding this patient resulted in MPFC NVLout beta value for controls vs. patients (p = 0.174) and POMS vs. MPFC NVLout beta values correlation (r = 0.08; p = 0.606). All beta values are shown as 10^−3^. Abbreviation: MPFC, medial prefrontal cortex; ROI, region of interest. POMS, Profile of Mood States.

In response to no-loss versus loss outcomes (NVLout), we found increased MPFC activity in patients but not controls. These results did not meet the corrected statistical threshold of p < 0.0125, but again, the trend was similar to the findings of Martucci et al. (2018) (Fig. 5). An ANCOVA analysis revealed that the covariate, age, was not significantly related to the MPFC NVLout fMRI beta values, F(1,37) = 2.007, p = 0.165. However, there was a significant group difference (which did not meet our pre-determined significance threshold) in the MPFC NVLout fMRI beta values after controlling for the effect of the age, F(1,37) = 4.418, p = 0.042.

Consistent with findings of Martucci et al. (2018), correlations of MPFC NVLout extracted beta values and mood disturbance (POMS), across combined patient and control groups, revealed a significant association (r = 0.45, p = 0.004). Post-hoc within-group analyses (i.e., correlations of POMS vs. ROI data from patients or controls) revealed a positive correlation only in the patient group (r = 0.582, p = 0.007). In Martucci et al. (2018), BSI depression and anxiety scales were not collected. After adding these scales, we found a significant correlation of MPFC NVLout extracted beta values and BSI anxiety across patient and control groups (r = 0.548, p < 0.001).Post-hoc within-group analyses identified a positive correlation only in the patient group (r = 0.659, p = 0.002). As described in the Methods, we included -$1 trials (in addition to -$5 trials) as an exploratory analysis to increase the power. The results did not meet the corrected statistical threshold, but again, suggested higher MPFC activity in the patient group relative to the control group (p=0.053; Supplementary Fig. 3).

For reward outcome responses (GVNout) and loss anticipation (LVNant), both groups showed increased MPFC BOLD signal for reward outcomes and reduced MPFC activity during loss anticipation, consistent with Martucci et al. (2018) (Supplementary Fig. 2).

### 3.6. Post-hoc fMRI Analysis

Post-hoc fMRI analyses were conducted on additional ROIs (VTA, aINS, ACC, and expanded MPFC) and for the whole brain. In Martucci et al. (2018), extracted beta values from the VTA, aINS, and ACC ROIs indicated slightly reduced activity in the patient group compared to controls during gain anticipation (GVNant). Our results did not replicate these patterns, with the exception that the bilateral aINS ROI showed reduced activity in the patient group relative to controls during GVNant (p = 0.04; Supplementary Fig. 4). As found in Martucci et al. (2018), the expanded MPFC mask analysis showed more extensive MPFC activation outside of our targeted MPFC ROI for both GVNant and NVLout contrasts (Supplementary Fig. 5). Lastly, a whole-brain analysis confirmed group differences in multiple brain regions (Supplementary Fig. 6).

## 4. DISCUSSION

The present study aimed to replicate the findings of Martucci et al. (2018) in a separate cohort of patients with fibromyalgia. In our pre-registered plan on the Open Science Framework (OSF, https://osf.io/4yctn), we hypothesized that patients with fibromyalgia would show altered brain reward activity compared to healthy controls in a way that patients would show altered MPFC activity in different rewarding situations but intact NAcc activity, as found in Martucci et al. (2018). Even though significance did not meet the pre-established threshold (consistent with Martucci et al., 2018; p < 0.0125), we found a similar trend of results. Specifically, patients showed reduced MPFC activity during reward anticipation and greater MPFC activity to punishment avoidance compared to controls. Also, consistent with the findings of Martucci et al. (2018), NAcc activity during reward anticipation and punishment avoidance did not differ between groups. We also replicated an individual difference correlation, such that patients with higher MPFC responses during punishment avoidance also reported greater total mood disturbance (POMS). Together, these findings generally support the notion of altered brain reward processing in individuals with fibromyalgia and replicate patterns of findings originally observed in Martucci et al (2018).

Correlational analyses of individual differences in clinical measures and extracted beta values of MPFC and NAcc in response to reward anticipation did not reveal significant associations. As all clinical measures (except the BIS scale) resemble those used by Martucci et al. (2018), extracted beta values might not be strong enough to resolve significant correlations. In Martucci et al. (2018), reduced reward anticipatory activity was observed in the ACC with a trend for a similar pattern in the aINS. We found reduced reward response in the aINS (p=0.04, uncorrected) but not in the ACC or VTA. However, in a whole-brain analysis that focused on a larger ACC region, the contrast activation map showed a similar pattern of results with Martucci et al. (2018), such that reduced reward anticipatory response in the ACC was observed in patients compared to controls (Supplementary Fig 4C).

Consistent with the findings of Martucci et al. (2018), MPFC activity in patients distinguished no-loss from loss outcomes (i.e., -$5 hit vs. miss) in patients to a greater extent than controls. Further, across controls and patients, the MPFC response to no-loss versus loss outcomes positively correlated with individual differences in mood disturbance (POMS). This result seemed due to the patient group since only within-group analysis in the patient group revealed a significant association (p = 0.007; controls: p = 0.458), supporting a prior finding of a relationship between negative expectations and altered MPFC response during loss avoidance in the patient group. A newly added clinical scale, BSI anxiety, also supported this interpretation by showing a positive association with the MPFC response to no-loss versus loss.

Even though the current results did not meet the predetermined significance threshold (p <0.0125), MPFC activity showed a trend towards similar patterns observed in Martucci et al. (2018). Specifically, the observed reduced MPFC response during gain anticipation and increased response during loss avoidance bolster previous findings and strongly support the notion of an altered brain reward response signature in patients with fibromyalgia. Overall, patients with fibromyalgia may process avoidance of punishment as more reinforcing than healthy controls, and even more reinforcing that obtaining rewards. As discussed previously, such a dysregulated MPFC BOLD response during reward processing may be related to altered midbrain dopamine function in fibromyalgia (Wood et al., 2007), altered prefrontal cognitive function (Glass, 2009; Seo et al., 2012), or a combination of the two. Additionally, we again observed relatively normal NAcc response during gain anticipation in patients (neither reduced as implied by (Wood et al., 2007) nor increased as implied by (Ledermann et al., 2017)). Consistent with our previous results, accentuated MPFC responses to nonreward outcomes were most characteristic of patients with fibromyalgia. Thus, this replication solidifies the importance of investigating further underlying related neurophysiologic, psychologic, and pharmacologic mechanisms which may inform improved therapeutics that target dysregulated brain reward function in fibromyalgia.

This replication study also has some limitations. First, findings did not fully replicate the findings of Martucci et al. (2018). All results showed a trend for similar patterns results, but none were significant at predetermined corrected thresholds. Several factors might have weakened the pattern of findings. While the procedure adhered as closely as possible to the original design, some minor deviations were apparent. While Martucci et al. (2018) used a Gradient Echo Pulse Sequence with “spiral in-out” acquisition, the current study used a Gradient Echo Pulse Sequence. As the spiral in-out scan sequence may improve the acquisition of medial prefrontal signal (Glover & Law, 2001), this different imaging acquisition technique may have influenced the results. Also, age significantly varied between groups in the current dataset. Although ANCOVA analyses revealed that the age covariate did not alter the main findings, still the average age of the patient group in the current study was younger than Martucci et al. (2018) (current study: M=35.9, SD=12.3; Martucci et al.: M = 48.1, SD = 9.6). Pain duration also differed slightly, which might have affected the results (current study: M = 6.03, SD = 5.33; Martucci et al.: M = 11.5, SD = 7.7). Lastly, for some of the participants (patients: N= 11; controls: N=16), since participation in the current study involved a second exposure to the MID task, their motivation might be changed as a function of repeated exposure. However, Srirangarajan et al. (2021) showed decent test-retest reliability in MID task signal, even when repeated in the same session.

## 5. CONCLUSIONS

Martucci et al. (2018) first reported dysfunctional reward processing in the MPFC during reward anticipation and in response to non-punishment outcomes in chronic pain, and more specifically in patients with fibromyalgia. Using predetermined hypotheses (OSF, https://osf.io/4yctn), we sought to replicate those findings. A consistent trend of similar findings was observed in the current study. Thus, the present replication findings support the need for more targeted future research on neural reward circuits, particularly focused on the MPFC, in patients with fibromyalgia. By focusing on replicated dysfunction of reward processing, future studies should have enhanced potential to identify robust and clinically-relevant neurophysiological alterations in chronic pain. Together with the present study, continued research along these avenues may shepherd a better understanding of the interaction between the experience of pain and dysregulated brain and behavior processes, as well as provide essential insights for improved treatments of chronic pain and comorbid affective conditions.

## Data Availability

All data produced in the present study are available upon reasonable request to the authors.

## Author Contributions

The authors confirm contribution to the paper as follows: S.H.P. and K.T.M. were responsible for the conception and design of the study. S.H.P., E.Z.D., A.K.B., and K.T.M. collected the data. S.H.P., E.Z.D., and K.T.M. analyzed the data. S.H.P., E.Z.D., A.K.B., K.H.M., B.K., and K.T.M. were involved in the interpretation of results. S.H.P. produced the initial manuscript draft. S.H.P., E.Z.D., A.K.B., K.H.M., B.K., and K.T.M. edited and revised the manuscript. All authors reviewed and approved the final version of the manuscript.

## Declarations of Conflict of Interest

The authors declare no competing interests.

## Acknowledgments

We thank Lindsie Boerger, Meghna Nanda, Vinit Krishna, and Sharon Norman for their assistance with recruitment, data collection, data organization, regulatory oversight, and assistance with data quality control. We also very sincerely thank the Duke University Brain Imaging and Analysis Center (BIAC), particularly, Drs. Allen Song and Todd Harshbarger, for assistance with fMRI sequence and protocol design, and Susan Music, Jennifer Graves, and Lamont Cannon for their expert assistance with MRI/fMRI data collection. Lastly, we thank all of the study participants for their time and contribution to advance clinical research and knowledge. For this study the authors received funding from the National Institutes of Health R00 DA040154 (K.T.M.) and the Stanford Wu Tsai Neuroscience Institutes’ Neurochoice Initiative (B.K.).

## Notes

### Competing Interest Statement

The authors have declared no competing interest.

### Funding Statement

This project was funded by the National Institutes of Health, National Institute of Drug Abuse (NIDA), R00 DA040154 (awarded to K.T.M.), and the Stanford Wu Tsai Neuroscience Institutes′ Neurochoice Initiative (B.K.).

### Author Declarations

All study procedures were approved by the Duke University Institutional Review Board.

