## Supplementary Figures for "Altered brain reward response to monetary incentives in fibromyalgia: A replication study"

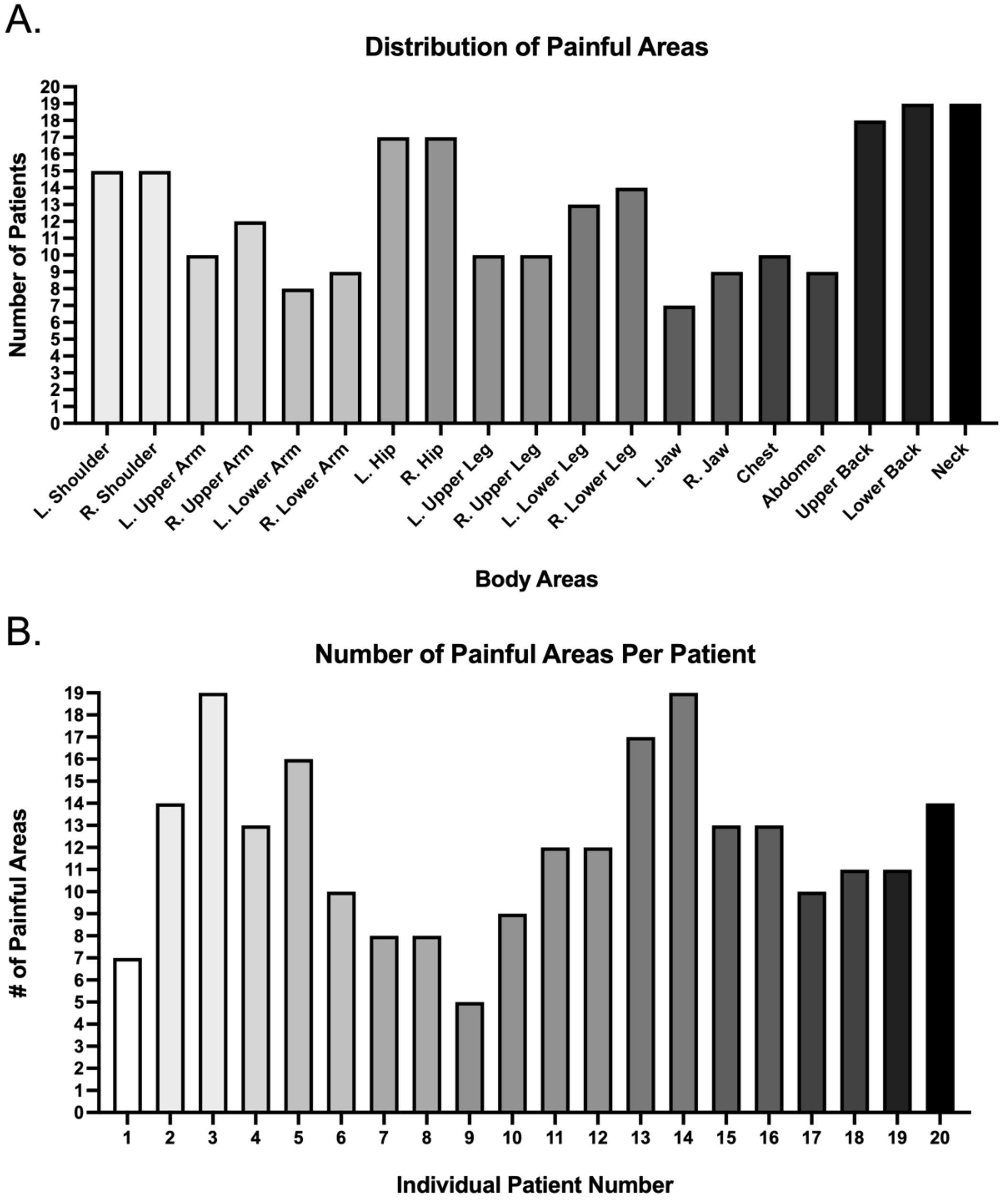


Supplementary Figure 1. Distribution and Number of Painful Areas in Patients.

(A) Based on the Fibromyalgia Assessment Form, patients were asked to report the presence or absence of pain in 19 body regions. The number of patients reporting pain in each area is shown in the y-axis (total N = 20 patients). B) The number of painful body areas reported by each patient (N = 20) is shown (maximum number of body regions = 19). In both figures, greyscale shaded bar colors are to increase the readability of results and do not represent additional data.

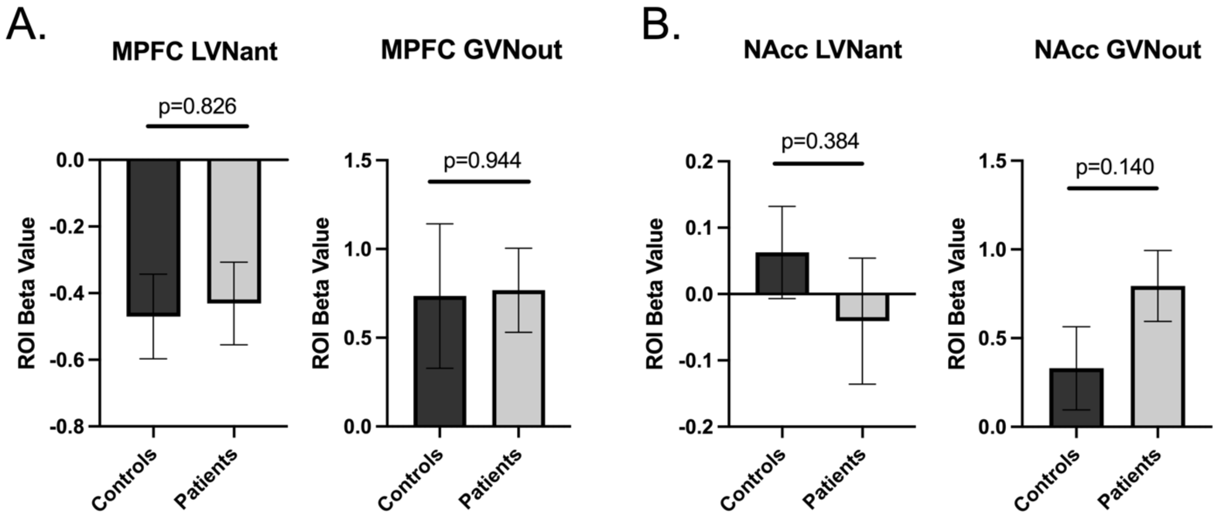


Supplementary Figure 2. MPFC and NAcc ROI Activity in Response to Loss vs. No-loss Anticipation and Gain vs. No-gain Outcomes.

(A) Contrast beta values extracted from the MPFC ROI in response to loss vs. no-loss anticipation (i.e., $0 vs. -$5 anticipation; NVLant) and gain vs. no-gain outcome (i.e., +$5 hit vs. miss; GVNout). (B) Contrast beta values extracted from the NAcc ROI in response to loss vs. no-loss anticipation (i.e., $0 vs. -$5 anticipation; NVLant) and gain vs. no-gain outcome (i.e., +$5 hit vs. miss; GVNout). Abbreviation: MPFC, medial prefrontal cortex; NAcc, nucleus accumbens; ROI, region of interest.

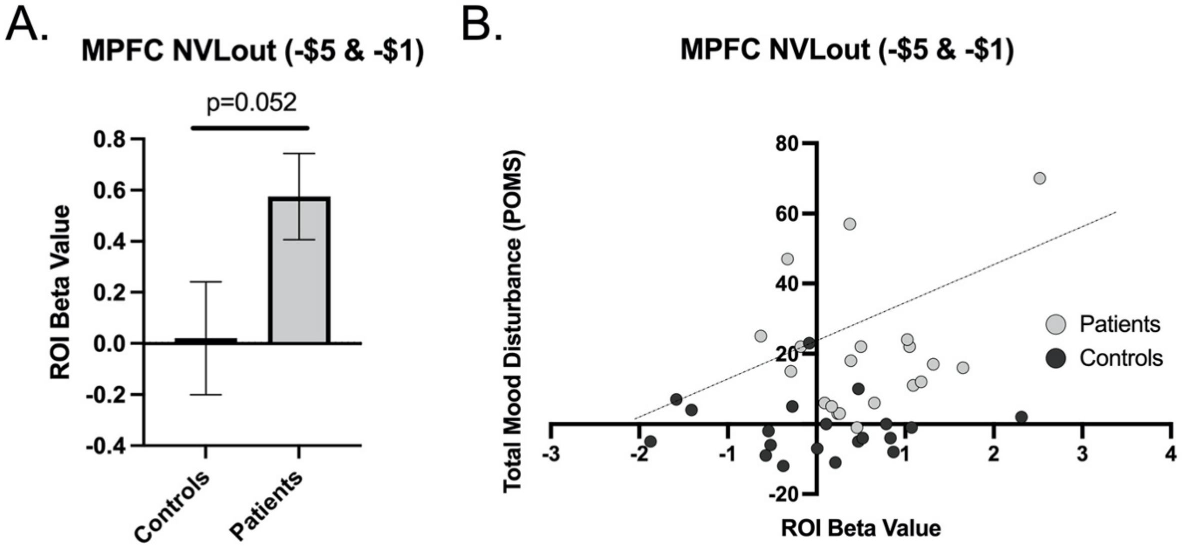


Supplementary Figure 3. MPFC ROI Activity in Response to No-loss vs. Loss outcomes.

(A) Contrast beta values extracted from the MPFC ROI during reward outcome (i.e., -$5 & -$1 hit > miss; NVLout). As compared to anticipation trials, the number of outcome trials in each condition (i.e., -$5 and -$1/hit and miss) varied, which might yield less power; therefore, as an exploratory analysis, we included -$1 trials to the NVLout condition to increase the number of trials included. (B) Correlations between MPFC NVLout (-$5 and -$1 outcome trials) extracted beta values and total mood disturbance (POMS) for both groups. One patient who showed a high MPFC NVLout beta value (see results in Fig. 5D) did not have as high beta value in the NVLout contrast with both -$5 and -$1 data. Post-hoc analyses resulted in trend significance for MPFC NVLout (-$5 & -$1) beta value for controls vs. patients ($p = 0.052$) and POMS vs. MPFC NVLout (-$5 & -$1) beta values correlation ($r = 0.297$; $p = 0.062$). All beta values are shown as $10^{-3}$. Abbreviation: MPFC, medial prefrontal cortex; ROI, region of interest. POMS, Profile of Mood States.

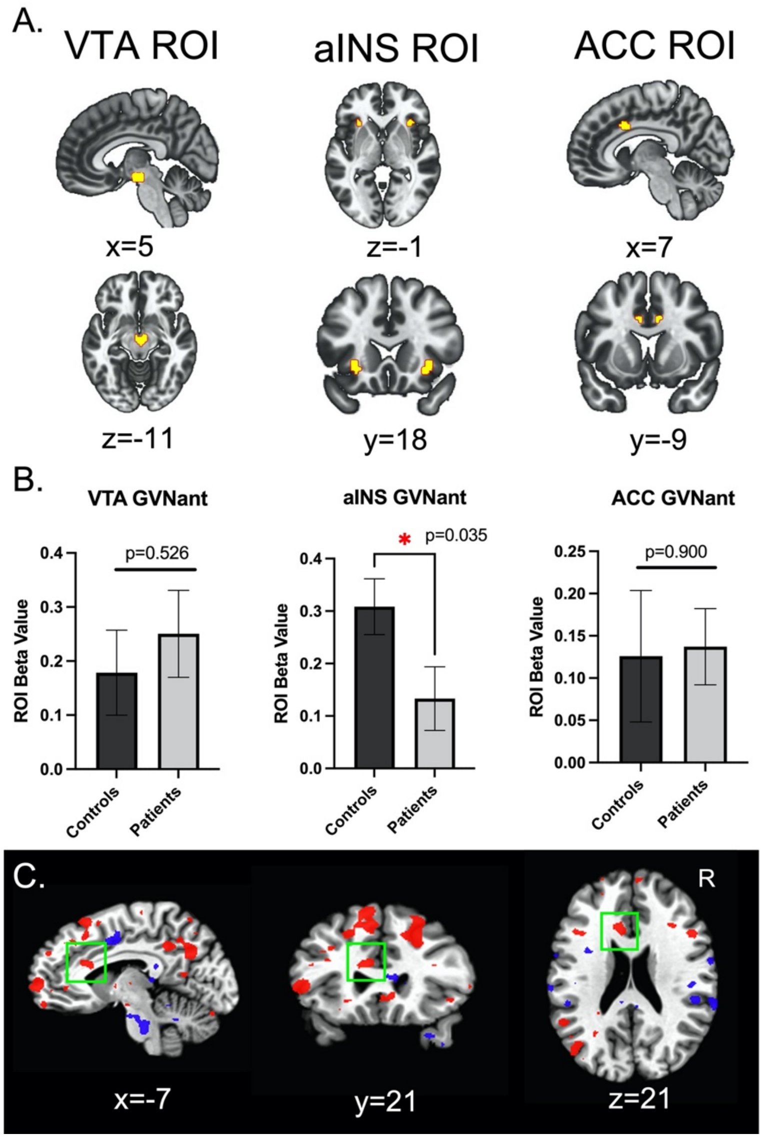


Supplementary Figure 4. Additional Regions of Interest during Reward Anticipation.

(A) ROIs of the ventral tegmental area (VTA), anterior insular cortex (aINS), and anterior cingulate cortex (ACC). (B) Contrast (+$5 vs. $0 anticipation, GVNant) extracted ROI beta values. These were post-hoc analyses, so they were not corrected for multiple comparisons. We found a significant difference between groups in the aINS. Martucci et al. (2018) did not show a group difference in the aINS, but the pattern of results is similar (i.e., higher mean beta value with controls compared to patients). (C) Inconsistent with Martucci et al. (2018), the ACC showed similar beta values between groups. We ran a whole-brain analysis to explore this discrepancy further and focused on a larger ACC region. As depicted in (C), contrast (controls vs. patients; $p < 0.05$, uncorrected; red color indicates positive value) activation map showed a similar pattern of nearby ACC activation as in Martucci et al. (2018). All beta values are shown as $10^{-3}$. Abbreviations: VTA, ventral tegmental area; aINS, anterior insular cortex; ACC, anterior cingulate cortex; R., right; ROI, region of interest.

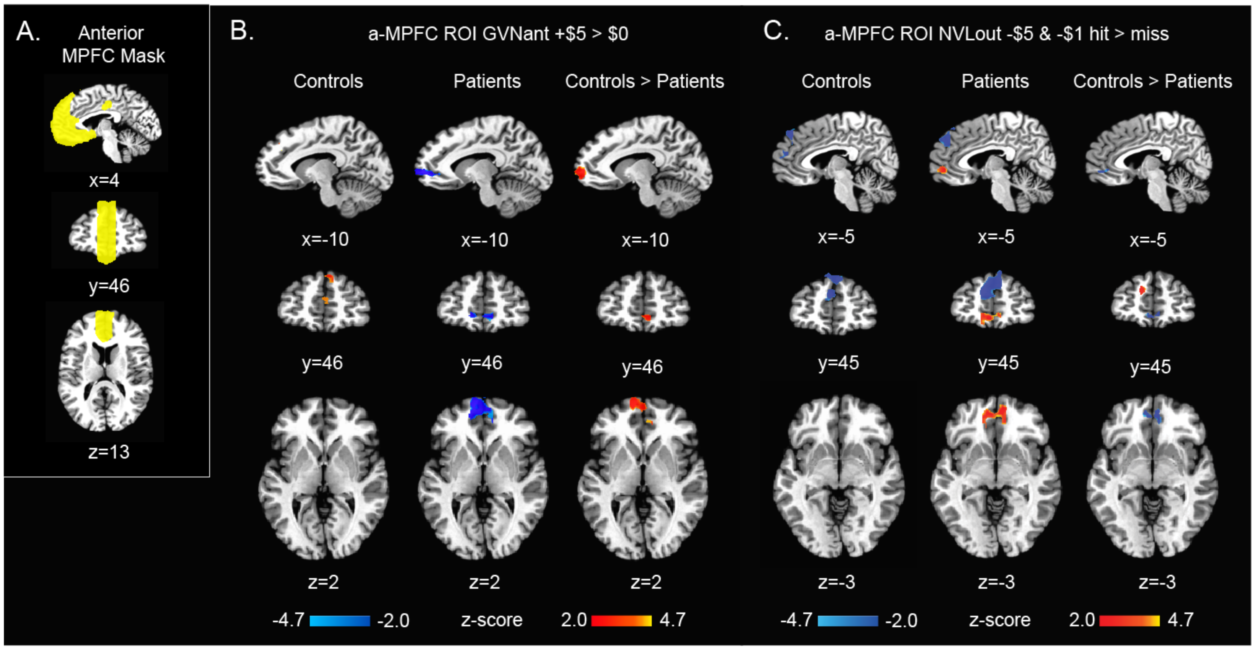


Supplementary Figure 5. Gain vs. No-gain Anticipation and No-Loss vs. Loss Outcome Activity within Anterior Medial Prefrontal Cortex Mask.

(A) The same expanded mask of the anterior medial prefrontal cortex (a-MPFC) that Martucci et al. (2018) used. (B) Activation maps showing control and patient groups and group differences of the a-MPFC activity for reward gain anticipation (GVNant) and (C) no-loss outcome (NVLout) contrasts ($p < 0.05$, uncorrected). Abbreviations: a-MPFC, anterior medial prefrontal cortex; ROI, region of interest.

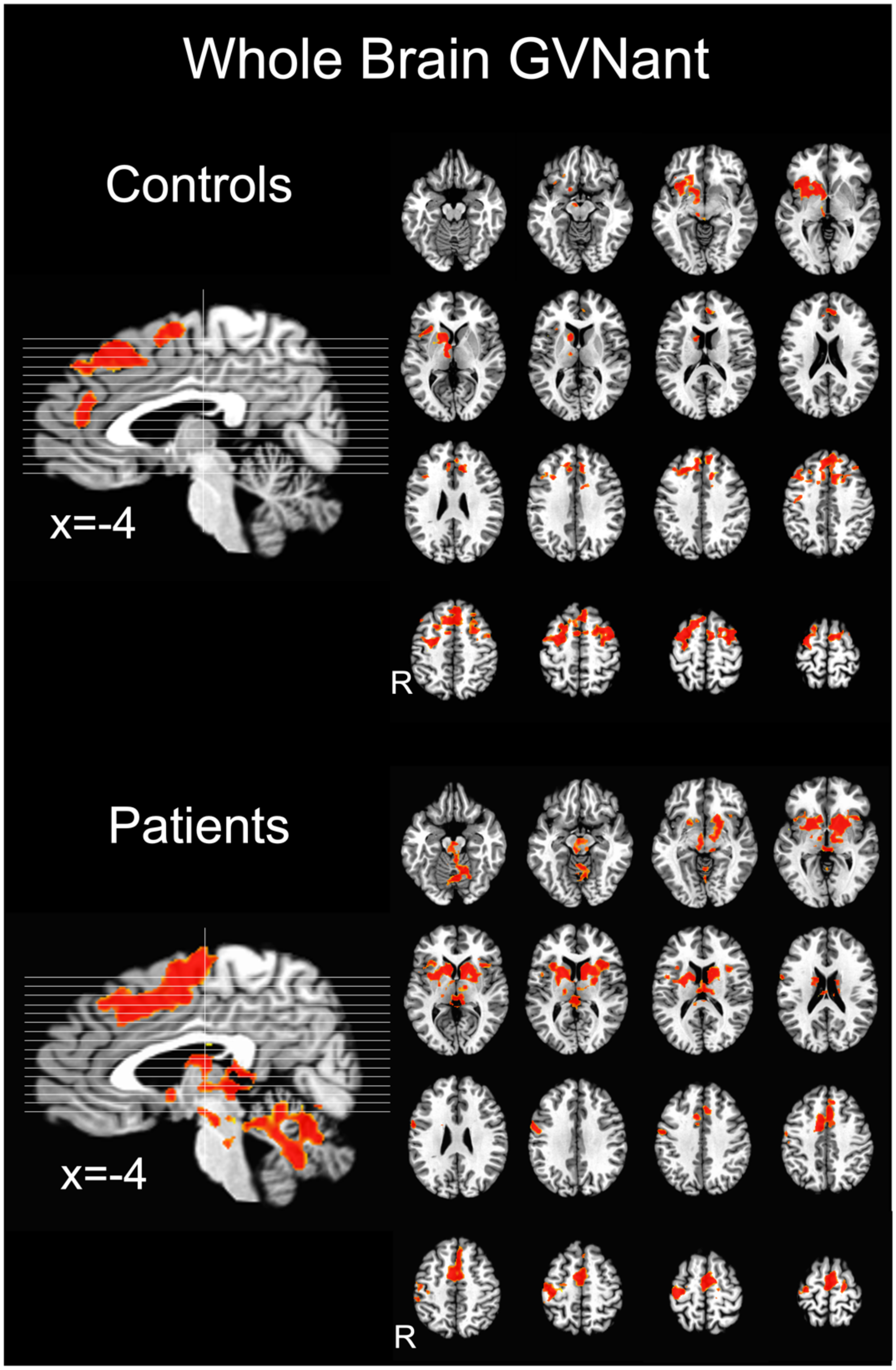


Supplementary Figure 6. Whole Brain Activation during Reward Gain Anticipation.

A post-hoc whole-brain analysis was conducted to compare our findings with Martucci et al. (2018) and confirm our main results. Contrast (+$5 vs. $0 anticipation, GVNant) activation maps are shown for the control group and patient group ($p < 0.05$, corrected alpha $< 0.05$, clusters that did not pass these thresholds are not presented). Abbreviation: R, right.
