## Supplementary Table 1 for "Altered brain reward response to monetary incentives in fibromyalgia: A replication study"

**Table S1. Clincial, Behavioral, and Psychological Correlations with ROI Betas**

| | | Patients and Healthy Controls | | | Patients Only | | | Controls Only | | |
|---|---|---|---|---|---|---|---|---|---|---|
| | | **NAcc GVNant** | **MPFC GVNant** | **MPFC NVLout** | **NAcc GVNant** | **MPFC GVNant** | **MPFC NVLout** | **NAcc GVNant** | **MPFC GVNant** | **MPFC NVLout** |
| **Positive Affect (PANAS)** | **r** | -0.01 | 0.232 | -0.194 | 0.147 | 0.142 | -0.236 | -0.088 | 0.044 | 0.119 |
| | **p** | 0.949 | 0.149 | 0.229 | 0.537 | 0.551 | 0.316 | 0.713 | 0.854 | 0.619 |
| | **N** | 40 | 40 | 40 | 20 | 20 | 20 | 20 | 20 | 20 |
| **Negative Affect (PANAS)** | **r** | -0.004 | -0.152 | 0.154 | -0.057 | -0.051 | 0.153 | 0.027 | -0.051 | -0.059 |
| | **p** | 0.983 | 0.348 | 0.341 | 0.811 | 0.832 | 0.521 | 0.909 | 0.831 | 0.805 |
| | **N** | 40 | 40 | 40 | 20 | 20 | 20 | 20 | 20 | 20 |
| **Mood Disturbance (POMS)** | **r** | -0.057 | -0.078 | **0.45****| -0.164 | 0.172 | **0.582**** | 0.033 | 0.07 | -0.176 |
| | **p** | 0.727 | 0.631 | **0.004** | 0.489 | 0.468 | **0.007** | 0.891 | 0.771 | 0.458 |
| | **N** | 40 | 40 | **40** | 20 | 20 | **20** | 20 | 20 | 20 |
| **BAS Drive (BIS/BAS)** | **r** | -0.152 | 0.302 | 0.123 | -0.031 | 0.332 | 0.219 | -0.236 | 0.244 | 0.112 |
| | **p** | 0.35 | 0.058 | 0.45 | 0.898 | 0.152 | 0.355 | 0.317 | 0.301 | 0.638 |
| | **N** | 40 | 40 | 40 | 20 | 20 | 20 | 20 | 20 | 20 |
| **BAS Fun (BIS/BAS)** | **r** | -0.239 | 0.321 | 0.203 | **-0.524** | **0.509*** | 0.193 | 0.009 | 0.08 | 0.366 |
| | **p** | 0.137 | 0.043 | 0.21 | **0.018*** | **0.022** | 0.416 | 0.969 | 0.738 | 0.112 |
| | **N** | 40 | 40 | 40 | **20** | **20** | 20 | 20 | 20 | 20 |
| **BAS Reward (BIS/BAS)** | **r** | -0.118 | 0.123 | 0.229 | -0.181 | 0.354 | 0.176 | -0.074 | -0.02 | 0.288 |
| | **p** | 0.467 | 0.451 | 0.155 | 0.444 | 0.126 | 0.459 | 0.757 | 0.934 | 0.218 |
| | **N** | 40 | 40 | 40 | 20 | 20 | 20 | 20 | 20 | 20 |
| **BIS Total (BIS/BAS)** | **r** | 0.285 | -0.378 | -0.04 | **0.496*** | -0.392 | -0.016 | 0.134 | -0.295 | -0.205 |
| | **p** | 0.074 | 0.016 | 0.808 | **0.026** | 0.087 | 0.945 | 0.574 | 0.207 | 0.386 |
| | **N** | 40 | 40 | 40 | **20** | 20 | 20 | 20 | 20 | 20 |
| **Depression (BDI)** | **r** | 0.119 | -0.143 | 0.053 | 0.321 | -0.093 | -0.029 | -0.208 | 0.113 | -0.34 |
| | **p** | 0.475 | 0.392 | 0.754 | 0.181 | 0.704 | 0.905 | 0.394 | 0.646 | 0.154 |
| | **N** | 38 | 38 | 38 | 19 | 19 | 19 | 19 | 19 | 19 |
| **State Anxiety** | **r** | 0.024 | -0.104 | 0.409 | -0.099 | 0.028 | **0.637*** | 0.129 | 0.009 | -0.16 |
| | **p** | 0.884 | 0.529 | 0.01 | 0.677 | 0.906 | **0.003** | 0.598 | 0.97 | 0.513 |
| | **N** | 39 | 39 | 39 | 20 | 20 | 20 | 19 | 19 | 19 |
| **Trait Anxiety** | **r** | 0.133 | -0.262 | 0.086 | 0.259 | -0.236 | 0.159 | 0.049 | -0.037 | -0.329 |
| | **p** | 0.418 | 0.108 | 0.604 | 0.284 | 0.33 | 0.515 | 0.836 | 0.878 | 0.157 |
| | **N** | 39 | 39 | 39 | 19 | 19 | 19 | 20 | 20 | 20 |
| **Pain Severity (BPI)** | **r** | | | | 0.225 | 0.365 | 0.073 | | | |
| | **p** | N/A | N/A | N/A | 0.339 | 0.113 | 0.76 | N/A | N/A | N/A |
| | **N** | | | | 20 | 20 | 20 | | | |
| **Pain Interference (BPI)** | **r** | | | | 0.043 | 0.027 | -0.1 | | | |
| | **p** | N/A | N/A | N/A | 0.858 | 0.909 | 0.674 | N/A | N/A | N/A |
| | **N** | | | | 20 | 20 | 20 | | | |
| **Fatigue (PROMIS)** | **r** | -0.012 | -0.216 | 0.083 | 0.167 | -0.09 | -0.127 | -0.206 | 0.143 | -0.405 |
| | **p** | 0.941 | 0.181 | 0.611 | 0.483 | 0.706 | 0.594 | 0.383 | 0.549 | 0.076 |
| | **N** | 40 | 40 | 40 | 20 | 20 | 20 | 20 | 20 | 20 |
| **BSI Depression** | **r** | -0.151 | -0.01 | 0.327 | -0.289 | 0.093 | **0.533*** | -0.072 | 0.249 | -0.29 |
| | **p** | 0.352 | 0.952 | 0.039 | 0.216 | 0.698 | **0.016** | 0.762 | 0.289 | 0.214 |
| | **N** | 40 | 40 | 40 | 20 | 20 | **20** | 20 | 20 | 20 |
| **BSI Anxiety** | **r** | -0.164 | -0.05 | **0.548***** | -0.333 | 0.24 | **0.659**** | 0.034 | -0.228 | 0.093 |
| | **p** | 0.31 | 0.757 | **< .001** | 0.151 | 0.308 | **0.002** | 0.886 | 0.334 | 0.696 |
| | **N** | 40 | 40 | **40** | 20 | 20 | **20** | 20 | 20 | 20 |

**Supplementary Table 1. Clinical, Behavioral, and Psychological Correlations with ROI Betas**. The total number of participants are 40 (patients N=20, controls N=20). Since not all participants completed BDI, State Anxiety, and State Trait, the total number of these measures differs from other measures. Abbreviations: PANAS, Positive and Negative Affect Schedule; POMS, Profile of Mood States; BAS/BIS, Behavioral Activation System/Behavioral Inhibition System; BDI, Beck Depression Inventory; STAI-State, STAI-Trait (State-Trait Anxiety Inventory); BPI, Brief Pain Inventory; PROMIS, Patient-Reported Outcomes Measurement Information System version Bank 1.0; BSI, Brief Symptom Inventory; p, P-value; r, Pearson correlation; N, number of participant data sets included in each analysis. Bold text indicates results meeting the significance criteria for between-group (patients and controls) correlations Bonferroni corrected for multiple comparisons $p < 0.008$, and for post-hoc associated within-group (patients or controls) correlations $p < 0.05$ (uncorrected). Red color text indicates a clinical measure that replicated resuls from Martucci. et al. (2018).
